# Genome-wide analysis of 439 mass spectrometry-based proteomic profiles in a population of 15,035 Scottish individuals

**DOI:** 10.1101/2025.08.14.25333677

**Authors:** Anne Richmond, Josephine A. Robertson, Hannah M. Smith, Robert F. Hillary, Aleksej Zelezniak, Spyros Vernardis, Aleksandra D. Chybowska, Arturas Grauslys, Jure Mur, Archie Campbell, Camilla Drake, Hannah Grant, Poppy Adkin, Matthew White, Charles Brigden, Christoph B. Messner, David J. Porteous, Caroline Hayward, Andrew M McIntosh, Daniel L. McCartney, Markus Ralser, Riccardo E. Marioni

## Abstract

Understanding the genetic architecture of the circulating proteome can help determine if a protein is causally linked to disease. Previous large-scale genome-wide association studies (GWAS) of proteins have mostly been conducted to pre-defined, targeted subsets of the proteome, and have often concentrated on low abundance proteins, many of which don’t exert their main function in serum. Mass spectrometry-based proteomics facilitates the study of high-abundance proteins and their isoforms, focussing on proteins active in blood. In 15,035 individuals from Generation Scotland, we performed GWAS of 439 highly abundant serum protein groups as identified and quantified by liquid chromatography tandem mass spectrometry. We identified 1,553 independent SNP signals for 398 proteins (P_Bonferroni_ < 1.2×10^−10^). Two-sample Mendelian Randomisation (MR) analyses were applied to test if the 398 proteins with significant SNP signals were causally associated with 79 common causes of morbidity and mortality. We report putative causal associations between 13 proteins and 17 outcomes including neuropsychiatric and cardiovascular conditions. Large scale genome-wide analyses of the high abundance proteome complement targeted approaches for the discovery of causal pathways of disease.

## Introduction

The circulating proteome can capture a readout of lifestyle and environmental exposures and inform disease risk prediction^1^. As a consequence, serum proteins are used as biomarkers in several conditions, such as cardiac Troponin I and T to indicate myocardial infarction^2^, cystatin C to indicate kidney dysfunction^3^, C-reactive protein to indicate inflammation^4^, or prostate specific antigen (PSA) as a marker of prostate cancer^5^. In addition to biomarker work, understanding the genetic architecture of blood-based proteins helps to determine causal disease mechanisms and identify drug targets^6^.

To date, large-scale studies have used targeted aptamer- or affinity-based platforms, that can assess upwards of 3,000 (Olink Explore 3,072 panel) and 7,000 pre-defined proteins (SomaLogic SomaScan assay)^7,8^. Genome wide association studies (GWAS) of these proteins have helped to give mechanistic insights into multiple disease outcomes – including those where blood is not the primary tissue of interest^9–12^. However, assay-based techniques have limitations. Quantification of functionally important, high abundance serum proteins can be inaccurate due to signals exceeding the dynamic range of the array and the presence of different isoforms^13^. They are also limited by a pre-defined list of protein targets, off target binding and interference caused by autoantibodies^14^, as well as lack of concordance across arrays^15^.

Untargeted approaches for proteomics such as mass spectrometry are unbiased and yield comprehensive and granular coverage of the high-abundance proteome, down to the peptide level. A small number of studies have performed GWAS on mass spectrometry-measured proteins. However, their sample sizes have been limited to under 3,000 individuals^16,17^. Recently, we and others have developed an ultra-high-throughput mass spectrometry proteomic technology that can measure proteins and peptides in excess of 200 samples per day, while advances in sample processing and data analysis have increasingly mitigated the impact of problematic batch effects^18^. The application of this technology to large population biobanks will accelerate insights into the genetic control of the human proteome by providing an unprecedented scale and accuracy of protein quantification.

Here, we characterise the genetic architecture of 439 mass spectrometry-measured proteins in 15,035 individuals from the Generation Scotland study. First, we identify genetic variants that associate with mean differences in serum protein levels (i.e. protein quantitative trait loci, or pQTLs). Second, harnessing these data, we perform Mendelian Randomisation (MR) to explore causal associations between protein levels and 79 leading causes of morbidity and mortality^19,20^. Together, we leverage unbiased protein measurements to provide a catalogue of genetic influences on serum protein levels as well as detail their causal associations with prevalent disease states.

## Methods

### Generation Scotland

Generation Scotland is a family-based cohort study of 23,960 volunteers, aged 17 – 99 at recruitment^21^. Recruitment occurred between 2006 to 2011 when volunteers attended a baseline clinic where they completed health and lifestyle questionnaires, provided samples for biochemistry and omics analyses, and gave consent for data linkage to their medical records. There were 5,573 families with a mean size of 4 members and 1,400 participants without relatives. The median age at baseline was 47 years and the sample was 59% female. In the 15,035 individuals with both genetic and protein data, the median age (range of 18-99 years) was 49 years and the sample was 58% female.

### Genotyping

Genotyping was carried out using one of two arrays - the Illumina HumanOmniExpressExome-8v1-2_A or HumanOmniExpressExome-8v1_A. Quality control steps have been reported previously^22^. Briefly, they included filtering for single-nucleotide polymorphisms (SNPs) with missingness (>2% across all individuals), minor allele frequency (MAF) >1% with Hardy-Weinberg *p*-value <1×10^6^. Samples were removed based on sex mismatches with the phenotypic database and on missingness (>2% across a SNP). Genetic outliers were identified as lying more than six standard deviations (SDs) beyond the mean on the first two principal components upon integration with the 1,000 Genomes population^23^. These individuals were removed, leaving a dataset of 604,858 SNPs and 20,032 individuals^24^. SNP data were then imputed using the Haplotype Reference Consortium (HRC) imputation panel^25^, yielding 24,161,581 variants with MAF >1% and INFO >0.4 for the downstream analyses.

### Proteomics

High-throughput mass spectrometry proteomics was carried out using our in-house developed method on serum samples pre-processed for protein denaturation and trypsinisation^26,27^. Liquid-chromatography-mass spectrometry (LC-MS) was carried out using a scanning SWATH method, analytical flow rate chromatography using the 1290 Infinity II system (Agilent) and a Triple TOF 6600 mass spectrometer (SCIEX)^18^. DIA-NN^28^ was used to process the output data using a human plasma spectral library^29^, with a precursor false discovery rate (FDR) set to 1%. Signals were annotated using UniProt^30,31^. Additional processing was carried out in R, including correction for within-batch drift^32^ and between-batch correction (limma v3.54.2)^33^. 439 proteins and protein groups were considered, 133 (30.3%) of these were uniquely mapped to one UniProt ID (termed “proteins”), with the remaining 306 (69.7%) mapping to multiple potential UniProt IDs (termed “protein groups”). 199 of these protein groups represent different isoforms of the same protein and 107 mixed peptides where the signal mapped to different proteins. In many cases, the groups with mixed labels contain related proteins, for example, Haptoglobin and Haptoglobin-related-protein for the protein group labelled P00739.P00738.H0Y300.J3KTC3.J3QR68.A0A0C4DGL8. Annotation of the 439 proteins considered in this study are provided in **Supplementary Table 1**.

### Statistical Analyses

#### pQTL discovery

Genome-wide association studies were used to identify pQTLs for protein levels (N=15,035, **Figure 1A**). Linear regression was run using the fastGWA command in GCTA^34^. Protein levels were transformed by rank-based inverse normalisation. Age, sex and 20 genetic principal components (to capture population structure) were included as fixed effect covariates in the GWAS models. Correlations between all protein levels and covariates are detailed in **Supplementary Table 2**. A sparse genetic relationship matrix (GRM) was generated using a GRM cut-off value of 0.05 and fitted as a random effect to control for family structure. A Bonferroni statistical significance threshold was set at P < 1.2×10^-10^ (genome-wide significance threshold divided by number of inferred proteins, 5×10^-8^/439). *Cis* pQTLs were defined as variants lying within 1Mb of the transcription start site for the gene encoding the primary protein target. Lead independent pQTLs were identified via conditional and joint analysis of the summary statistics for each protein using GCTA-COJO^34,35^. Default settings were specified, which included a stepwise selection procedure across 10Mb regions and SNP linkage disequilibrium (LD) cut-offs set to *R*^2^ < 0.9 to avoid multi-collinearity. The statistical significance threshold was once again set to P < 1.2×10^-10^. HRC^25^ imputed data from Generation Scotland was used as the reference panel for LD calculations.

**Figure 1.**
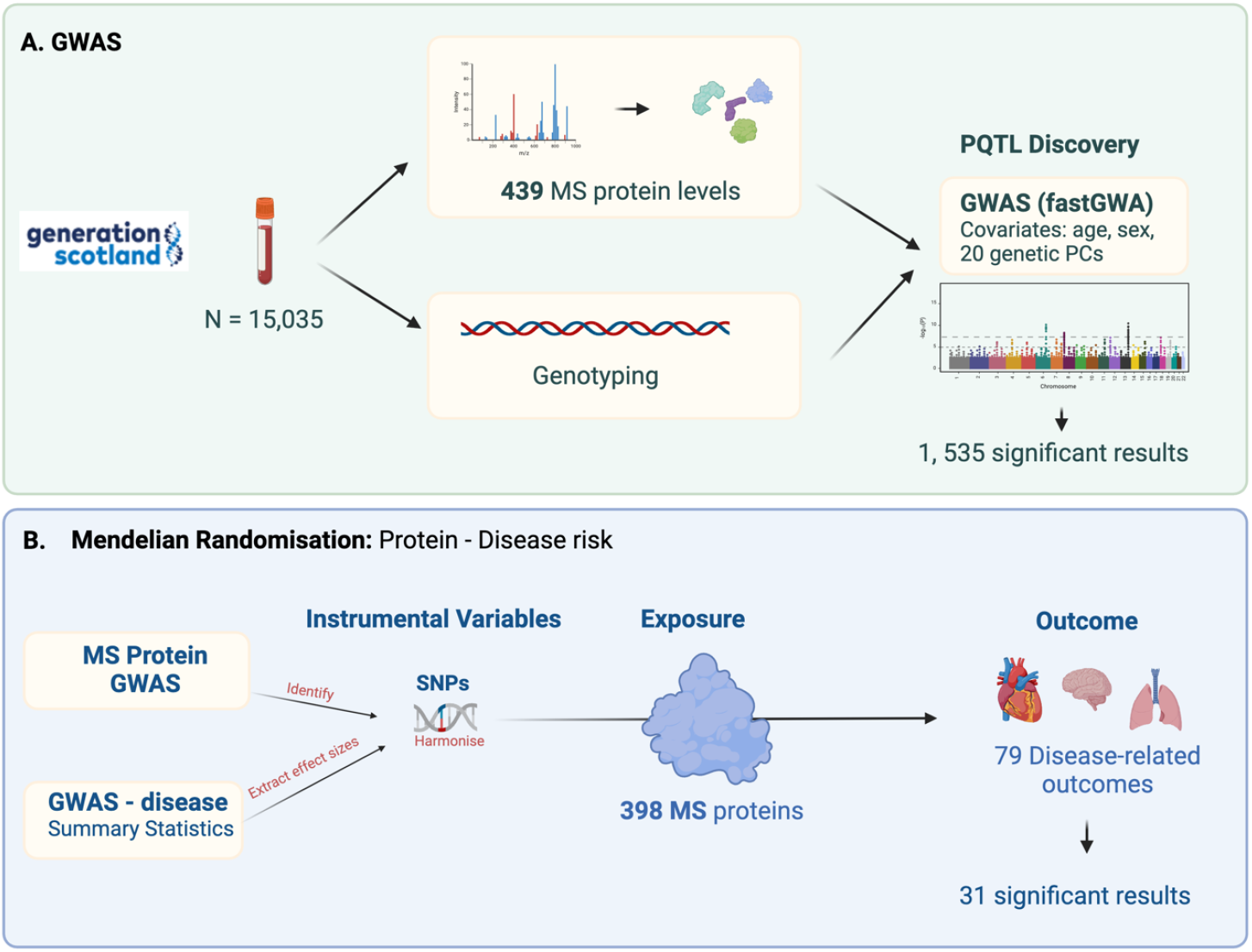
Outline of methods for GWAS and 2-sample MR. **(A) GWAS:** GWAS was run on 15,035 samples to identify pQTLs for 439 mass spectrometry proteins using the fastGWA option from GCTA. Age, sex and the first 20 genetic principal components were included as covariates. **(B) MR:** two-sample mendelian randomisation was carried out using results from our protein GWAS and summary statistics for GWASs of the 79 selected disease outcomes. MR was run for the 398 proteins with pQTLs in the GWAS results. Figure created with BioRender.com.

#### Comparison with previous pQTL studies

The discovered pQTLs were compared with those found in two previous large-scale pQTL studies by Sun *et al*^10^ and Eldjarn *et al*^9^. Sun *et al* used the Olink proximity extension assay for protein measurement in 54,265 UK biobank participants, while Eldjarn *et al* used the SomaScan v4 assay with a cohort of 35,892 Icelanders. Of the 439 proteins and protein groups quantified in our study, 58 and 75 were also included in the Sun *et al* and Eldjarn *et al* studies respectively. A pQTL was considered replicated if the genome-wide significant lead variant from the published studies was within 1Mb and with an effect size in the same direction as the discovered lead variant, as well as having an LD score of >0.6.

#### SNP-by-sex interactions

Sex differences in genetic effects have been described across complex traits^36^ but sex differences for molecular phenotypes remain largely underexplored. However, recently, a study of high-abundant plasma proteins measured by the same mass-spectrometry method as in our study, identified modulation of the proteome by hormonal contraceptive use^37^. To explore whether there may be sex-specific effects of SNPs on protein abundance, we repeated the primary pQTL analyses using fastGWA-GE^38^ with a SNP-by-sex interaction term for all proteins. A sparse GRM was specified as per the primary pQTL analyses, and the same stringent Bonferroni correction was applied.

#### Mendelian Randomisation

To investigate possible causal associations between protein levels and disease risk, two-sample MR analyses were carried out for each protein with >1 independent pQTL (exposure, n = 398) and a list of 78 common causes of morbidity and mortality taken from Gudjonsson *et al*.,^39^ plus critical COVID^40^ as outcomes (**Figure 1B**). A full list of these outcomes can be found in **Supplementary Table 3**, along with source information for the summary statistics. The lead independent SNPs were included as instruments for each protein and inverse-variance weighted (IVW) MR was run alongside MR-Egger, weighted median MR, weighted mode MR and simple mode MR sensitivity analyses. Evidence of directional horizontal pleiotropy was queried for by testing for a non-zero Egger MR regression intercept. MR Steiger directionality was used to determine the causal direction of the instruments on the exposure and outcome. A Bonferroni statistical significance threshold was set at P < 1.59 x 10^-6^ (0.05 /(number of outcomes x number of exposures)). An MR result was considered significant if the IVW p-value was below this threshold, at least one other method had P < 0.05 and it had at least 3 SNP instruments.

## Results

### pQTL discovery

GWAS analyses of 439 serum proteins (mapped to one UniProt ID) or protein groups (mapped to multiple UniProt IDs) levels were conducted using data from 15,035 participants from Generation Scotland. We observed 199,489 genome-wide significant (P_Bonferroni_ < 1.2×10^-10^) SNP associations with 398 (91%) of the proteins/protein groups. Following conditional analysis, 1,553 independent signals were identified with the lead variant defined as the SNP with the lowest P-value within the given locus (**Supplementary Table 4**). 1,199 of the significant pQTLs were found in uniquely mapped proteins (i.e, non-protein groups), 656 (54.7%) of which were *cis* (±1Mb), and 543 (45.3%) *trans* (**Figure 2A**). Over 70% of discovered pQTLs (404 *cis* and 200 trans) were associated with a single protein. Our results also indicated the presence of a pQTL hotspot within *SERPINA1*, with 17 independent signals generating 181 associations with 140 proteins. 71 proteins/protein groups had only one independent pQTL whilst 48 protein/protein groups had seven or more (**Figure 2B**). When comparing effect size with minor allele frequency, a negative correlation (for *cis* loci: r_Pearson_=-0.60, P=1.0×10^-64^; for *trans loci*: r_Pearson_= -0.72, P=3.6×10^-88^) between the log beta and the log MAF was observed (**Figure 2C**). In general, for the same MAF, larger effect sizes were found with *cis* loci than with *trans* (**Figure 2C**). An inverse association (r_Pearson_=-0.15, P=1.8×10^-4^) was found between strength of *cis* pQTL and distance from transcription start site (**Figure 2D**). The pQTL analysis was repeated to include a SNP-sex interaction term to test for sex differences in genetic effects. One significant association (P < 1.2×10^-10^) was found; a *cis* locus in Ceruloplasmin (CP) (**Supplementary Table 5**). This locus remained significant after running a sensitivity analysis which included contraception as a covariate to determine if the signal was being driven by hormonal contraceptive use.

**Figure 2.**
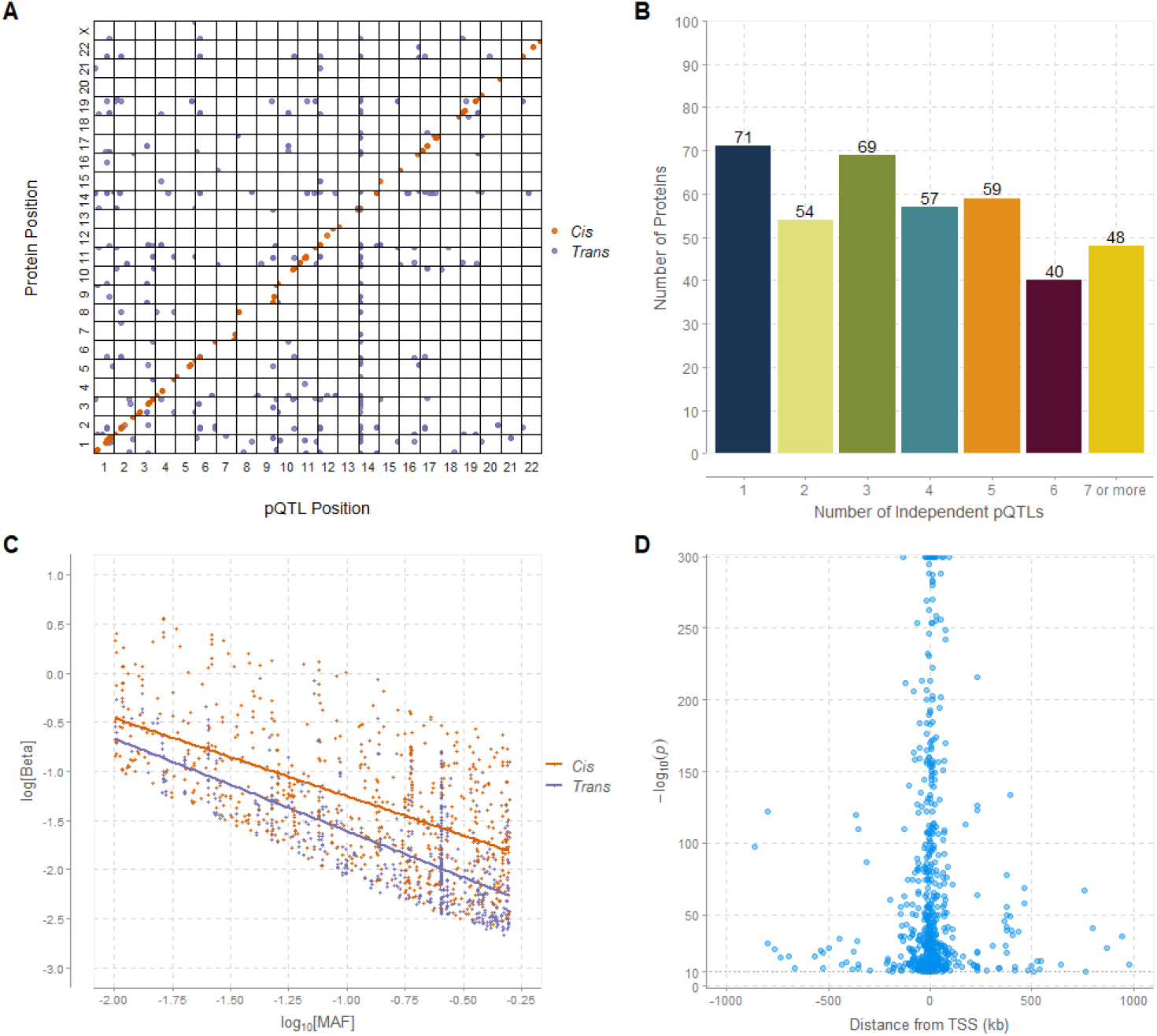
Genome-wide association studies to identify pQTLs for highly abundant proteins as measured by mass spectrometry. **(A)** The x-axis represents the chromosomal location of independent *cis* and *trans* pQTLs. The y-axis represents the position of the gene encoding the protein. Only uniquely mapped proteins are shown. *Cis* associations for proteins are marked by orange circles and *trans* associations (not linked to any protein targets) are marked by purple circles. **(B)** The number of independent pQTLs per protein (or protein group). **(C)** Association between the common logarithm of minor allele frequencies and the natural logarithm of absolute effect sizes for *cis* and *trans* pQTLs. Only uniquely mapped proteins are shown. *Cis* (orange circles and line); *trans* (purple circles and line). **(D)** Association between the strength of *cis* pQTL associations and their distance in kilobases (kb) from their respective transcription start sites. pQTL, protein quantitative trait locus; kb, kilobases.

### Replication of known genetic associations

The significant pQTLs discovered in our study were compared to two previous pQTL studies by Sun *et al*^10^ and Eldjarn *et al*.^9^ 51 and 69 proteins out of 398 with a significant association from our study overlapped with those in the Sun *et al* and Eldjarn *et al* studies, respectively. 46 associations out of 199 (23.12%) and 64 out of 275 (23.27%) found to be replicated in the two studies, respectively. A list of the overlapping proteins-SNP associations can be found in **Supplementary Tables 6 and 7**.

### MR of protein levels and 79 disease and health outcomes

Identifying causal protein – disease pathways could aid the discovery of novel drug targets; human genetic evidence increases the success rate for approval of developed drugs^41,42^. Given this, we tested whether any of the proteins or protein groups were causally associated with 79 disease or health outcomes (for full list see **Supplementary Table 3**). We identified 13 proteins that were putatively causal factors for 17 traits (**Supplementary Table 8**), giving a total of 31 significant results (inverse variance weighted (IVW) method with P_Bonferroni_ < 1.59 × 10^-6^, at least one other method with P < 0.05 and a minimum of 3 SNP instruments). The causal direction was from exposure (protein) to disease outcome, as determined by the Steiger directionality test. The proteins apolipoprotein E (APOE) and apolipoprotein B (APOB) were each associated with five of the cardiovascular related and metabolic traits (**Figure 3**). The strongest APOE association was with systolic blood pressure (IVW beta: 0.42, P: 8.5×10^- 16^) while the strongest APOB association was with LDL cholesterol (IVW beta: 1.17, P: 1.4×10^- 39^). Significant MR results were also found for three neuropsychiatric traits; autism spectrum disorder (ASD), Schizophrenia and Alzheimer’s disease (AD). The trypsin inhibitors inter-alpha-trypsin inhibitor heavy chain 3 (ITIH3) and inter-alpha-trypsin inhibitor heavy chain 4 (ITIH4) were associated with Schizophrenia (IVW log odds: -0.16, P: 1.7×10^-9^) and ASD (IVW log odds: -0.21, P: 6.3×10^-7^), respectively. Alpha-1-Antitrypsin (AAT) was also found to be associated with ASD (IVW log odds: -0.11, P: 4.2×10^-7^). AAT, ITIH3 and ITIH4 were also found to have associations or interactions with each other as defined by StringDB^43^ (**Supplementary Figure 1**). One protein, Immunoglobulin heavy constant mu (IGHM), showed evidence for a causal association with Alzheimer’s Disease (IVW log odds: -0.063, P = 4.3×10^-7^) (**Figure 4**).

**Figure 3.**
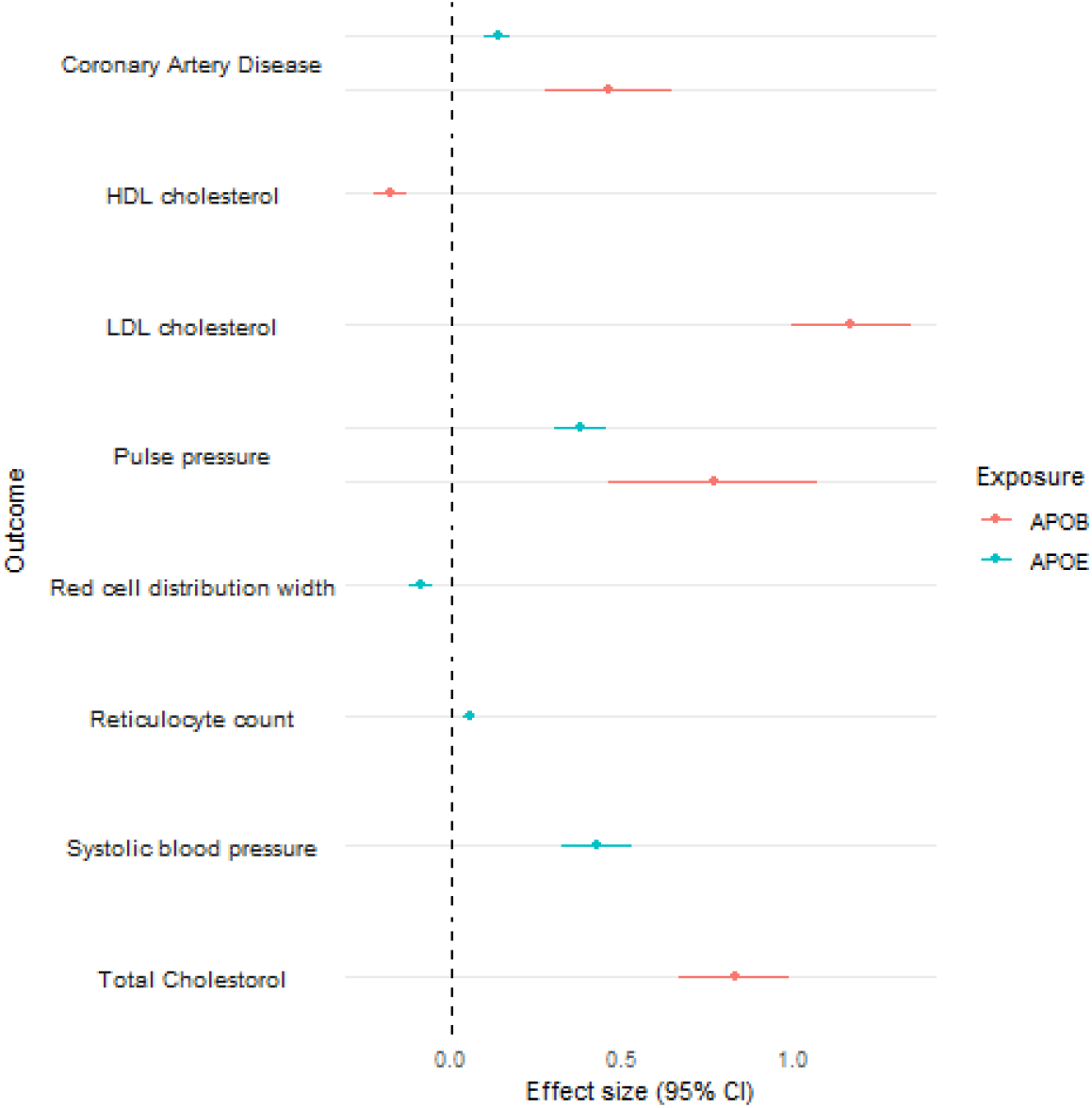
Traits associated with apolipoprotein B (APOB) apolipoprotein E (APOE) in MR analysis. Effect size and standard errors of the significant (P_Bonferroni_ <1.59×10^-6^) causal associations of APOB and APOE with 8 traits following MR analysis. Effect sizes displayed are log odds for binary traits and beta values for continuous traits, per SD of the rank transformed protein. Where multiple isoforms of the same protein were associated with one trait, the association with the largest effect size is shown.

**Figure 4.**
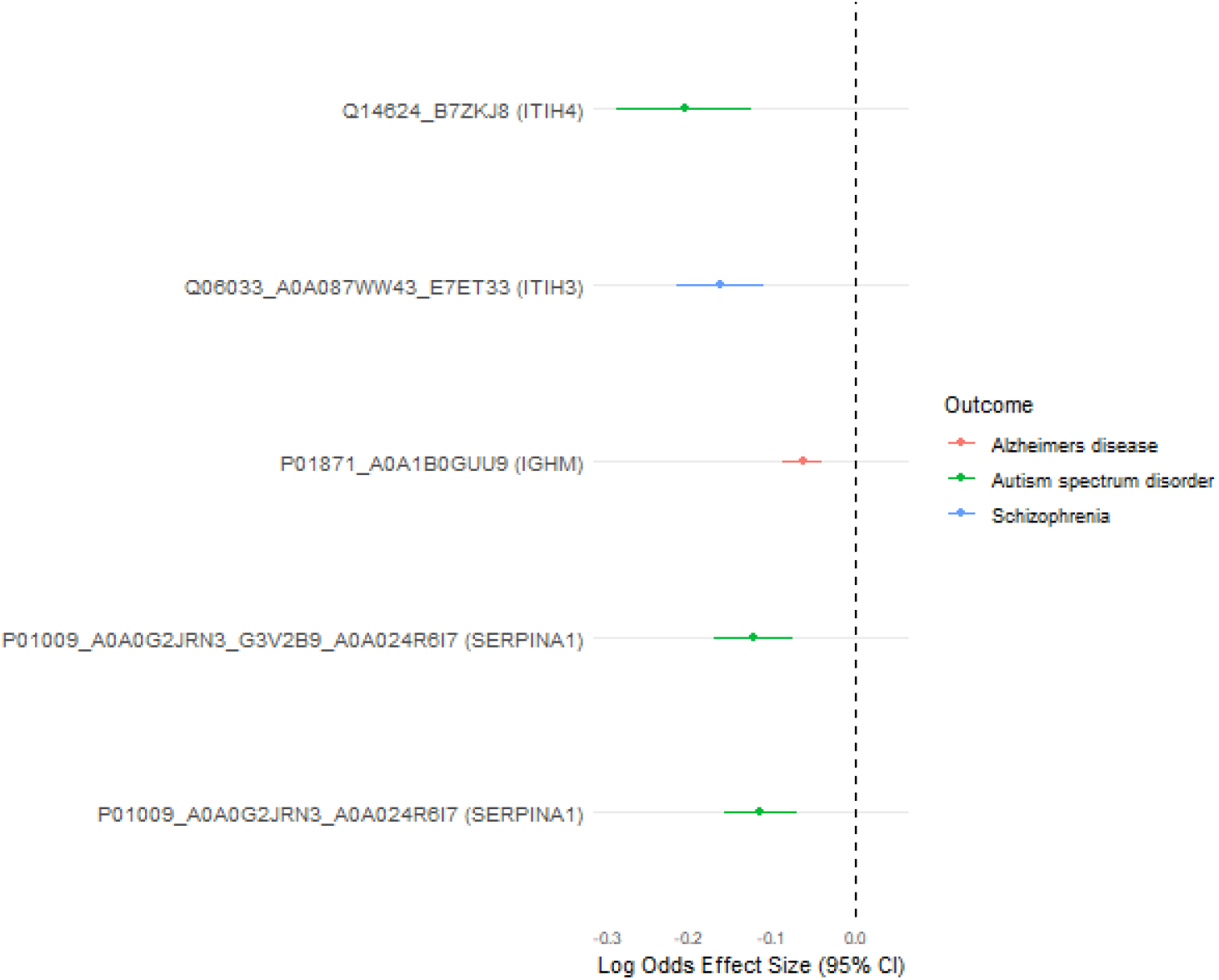
Proteins causally associated with Alzheimer’s disease (AD), Autism Spectrum Disorder (ASD) and Schizophrenia in MR analysis. Log odds effect size and standard errors of the significantly (P_Bonferroni_ <1.59×10^-6^) associated proteins with the neuropsychiatric conditions AD, ASD and Schizophrenia following MR analysis. Effect sizes are per SD of the rank transformed proteins.

## Discussion

Here, we describe a large-scale GWAS of 439 serum protein/protein group levels from the highly-abundant proteome fraction in the Generation Scotland cohort (N_samples_ = 15,035). The high-abundance proteome fraction is important for human physiology as it contains proteins important for metabolism, nutrient transport and the innate immune system including complement and coagulation factors. However, these proteins can be challenging to quantify as they exist in many isoforms and can saturate the measurement reagents. We identified 1,553 significant (P_Bonferroni_ < 1.2×10^-10^) independent pQTLs with 398 of the proteins/protein groups. Of the 1,199 loci significantly associated with unique proteins, we reported 656 as *cis* and 543 as *trans*. A *trans* hotspot was found within *SERPINA1*, with at least one significant association found with 140 of the 439 proteins. SERPINA1 encodes a serine protease inhibitor, AAT, predominantly produced in the liver^44^. SERPINA1 is involved in many biological processes, including inflammation, coagulation, homeostasis and the immune response^45^. Mutations in *SERPINA1* can result in AAT deficiency, increasing risk of liver and lung disease^44,46^. Given its involvement in a diverse range of pathways, it is perhaps unsurprising that pQTLs within *SERPINA1* have wide-ranging associations.

Whilst research into biomarkers for disease has identified many protein – disease associations, unpicking the biological mechanisms or causality for these associations is more challenging. MR analysis offers one route to assess for putative causal relationships between associations. We used two-sample MR to investigate causal associations between any of the proteins with identified pQTLs and 79 disease/health outcomes. There was evidence for causal associations between 13 proteins and 17 traits, including several neuropsychiatric disorders and cardiovascular-related traits.

MR analysis identified causal associations between APOB and APOE and coronary artery disease (CAD) and pulse pressure. Circulating APOB concentration is considered a biomarker for atherogenic lipids that are known to promote fatty deposits in arteries^47^. Polymorphisms in the APOE e4 allele have been shown to increase the risk of CAD^48^.

MR also revealed putative causal associations between anti-inflammatory proteins and neuropsychiatric disorders. Evidence is building for a role of immune-dysregulation in the aetiology of ASD and schizophrenia^49,50^. In our analysis, AAT and ITIH4 associated with ASD, and ITIH3 associated with schizophrenia, with all associations suggesting that increased protein levels reduce the odds ratio of diagnosis. These proteins have both anti-inflammatory and immunomodulatory functions^51^, have been implicated in the pathogenesis of autoimmune conditions and have also been proposed as potential therapeutic tools to manage these conditions^52^. They are produced both peripherally and in nervous-system tissue^43^. Our results build on existing evidence of a role of inter-alpha-trypsins in neuropsychiatric disorders from both genetic and proteomic studies. Schizophrenia is a highly heritable condition^53^ and previous large-scale GWAS studies have identified SNPs in *ITIH3/4* in association with Schizophrenia^54–56^ and also between *SERPINA1* and ASD^57^ and *ITIH3/4* and ASD^58^.

A few small-scale studies have identified associations between AAT and ASD. One study, using mass spectrometry proteomics identified increased expression level of AAT in participants with ASD (N_cases_ = 30, N_controls_ = 30)^59^, similarly another study using mass-spectrometry proteomics (N_cases_ = 30, N_controls_ = 30) identified increased AAT in children with ASD^60^. However, our results suggest that increased AAT could be protective against ASD.

Associations between AAT and Schizophrenia have also been demonstrated^61^, though our analysis did not identify a significant causal association. Our results support the hypothesis that AAT may perform an immunomodulatory role, which is protective against ASD. However, given that we have identified *SERPINA1* as a trans-hotspot for associations with multiple other proteins, including overlap in the SNP associations (see **Supplementary Table 4** and **Supplementary Table 8**), it is possible that genetic variants may affect the disease outcome through alternate pathways despite a non-zero Egger MR regression intercept.

Interestingly, we did not identify significant associations with Schizophrenia for all protein or protein groups mapped to AAT (total in the dataset = 5), though effect directions were concordant. These different entries may reflect different isoforms of the protein, which is known to have complex regulation and post-translational modifications^52^. Our findings of SERPINA1 as a *trans* pQTL hotspot further reinforces the complexity of its involvement in diverse molecular pathways. Mass spectrometry proteomics may prove to be a useful tool for studying similarly complex proteins, aiding the identification and study of different isoforms.

Another neurological condition with which we found a significant association is Alzheimer’s disease (AD). A causal relationship was found via MR with immunoglobulin heavy constant mu (IGHM), a component of immunoglobulin M (IgM), an antibody involved in the adaptive immune response and a function of which is to target and clear amyloid beta deposits. The immune system is thought to play a role in the progression of AD^63,64^ and decreased levels of IgM have been found in patients with AD than with age and sex matched healthy controls (n_cases_ = 26, n_controls_ = 26; n_cases_ = 30, n_controls_ = 30)^65,66^. However, findings on this topic have been inconsistent as IGHM has previously been shown to be a potential biomarker for AD, with increased levels found in patients with AD than with healthy controls (n_cases_ = 45, n_controls_ = 45)^67^. IgM levels in frontal cortex tissue have also been found to be increased in people with late-stage (stages V and VI) AD compared to healthy controls of the same age (n_cases_ = 11, n_controls_ = 9)^67,68^.

Technological advances in mass-spectrometry are increasing its potential for application both in large-cohorts and clinical settings^69^. Its untargeted approach avoids bias associated with antibody- or aptamer-based protein panels. Further, mass spectrometry best captures the high-abundance serum proteins^14^, which are not well quantified by assays reliant on an epitope, due to saturation and the presence of multiple isoforms^13,70^. Isoforms which modify protein affinity to assay epitopes, can interfere with MR analyses dependent on these results. Mass spectrometry is robust to isoform effects and therefore an important technique for identification or verification of disease-relevant pQTLs^71^, in addition to allowing exploration of high-abundance proteins less well captured by assay-based methods^72^. These high-abundance proteins are typically involved in processes occurring within the blood, such as nutrient transport, coagulation and innate immunity and have been less-prioritised in proteomic biomarker work thus far^73^. By concentrating on the highly abundant serum proteome fraction, we have not investigated lower-abundance proteins, predominantly associated with cell signalling or leakage from tissues^74^ which can yield meaningful biological insights^75^. This study benefits from a large sample size (N = 15,035) for pQTL analyses, giving us the ability to detect small effects. Our replication analysis was limited as protein GWASs for some of the proteins/peptides in our study do not exist. Nonetheless, our findings showed some concordance with those where replication was possible. This is encouraging given that different methods were used for protein quantification.

## Conclusion

We used a recently developed high-throughput mass spectrometry platform for a large-scale GWAS study of high-abundant circulating proteins, many of which function in plasma. Following pQTL identification, conditional and joint analysis revealed 1,553 significant and independent pQTLs for 398 proteins or protein groups, including a *trans* pQTL hotspot in *SERPINA1*. MR analysis revealed 13 proteins that have potential causal associations with 17 disease traits. Within these results, we found associations between trypsin inhibitors and the neuropsychiatric conditions ASD and schizophrenia. This demonstrates that the combined approaches of GWAS and MR can reveal biologically relevant protein networks which warrant further investigation for understanding of disease.

## Data Availability

According to the terms of consent for Generation Scotland participants, access to data must be reviewed by the Generation Scotland Access Committee. Applications should be made to. All code associated with this manuscript is available open access at the following GitHub repository: https://github.com/marioni-group/Generation_Scotland_Protein_GWAS. GWAS summary statistics are available on request and will be submitted to the Edinburgh DataShare site and GWAS Catalog upon publication.

## Acknowledgements

This research was funded in whole, or in part, by the Wellcome Trust [104036/Z/14/Z, 108890/Z/15/Z, 221890/Z/20/Z, 220857/Z/20/Z and 218493/Z/19/Z]. For the purpose of open access, the author has applied a CC BY public copyright licence to any Author Accepted Manuscript version arising from this submission.

GS received core support from the Chief Scientist Office of the Scottish Government Health Directorates [**CZD/16/6**] and the Scottish Funding Council [**HR03006**]. Genotyping of the GS samples was carried out by the Genetics Core Laboratory at the Edinburgh Clinical Research Facility, Edinburgh, Scotland and was funded by the Medical Research Council UK and the Wellcome Trust (Wellcome Trust Strategic Award “STratifying Resilience and Depression Longitudinally” (STRADL) Reference 104036/Z/14/Z). R.F.H. is supported by a British Heart Foundation Immediate Fellowship [**FS/IPBSRF/22/27042**]. C.H. is supported by an MRC Human Genetics Unit programme grant ‘Quantitative traits in health and disease’ [U. **MC_UU_00007/10**]. R.E.M. is supported by an Alzheimer’s Society major project grant **[AS-PG-19b-010**]. H.M.S is a student on the Translational Neuroscience PhD programme funded by Wellcome [**21843/Z/19/Z]**. J.A.R. is a University of Edinburgh Clinical Academic Track PhD student, supported by the Wellcome Trust [319878/Z/24/Z]. A.D.C. is supported by a Medical Research Council PhD Studentship in Precision Medicine with funding from the Medical Research Council Doctoral Training Program and the University of Edinburgh College of Medicine and Veterinary Medicine.

## Ethics statements

### Generation Scotland

All components of Generation Scotland received ethical approval from the NHS Tayside Committee on Medical Research Ethics [REC Reference Number: **05/S1401/89**]. Generation Scotland has also been granted Research Tissue Bank status by the East of Scotland Research Ethics Service [REC Reference Number: **20-ES-0021**], providing generic ethical approval for a wide range of uses within medical research. All participants provided written informed consent.

