## Supplementary Figure 1 for "Genome-wide analysis of 439 mass spectrometry-based proteomic profiles in a population of 15,035 Scottish individuals"

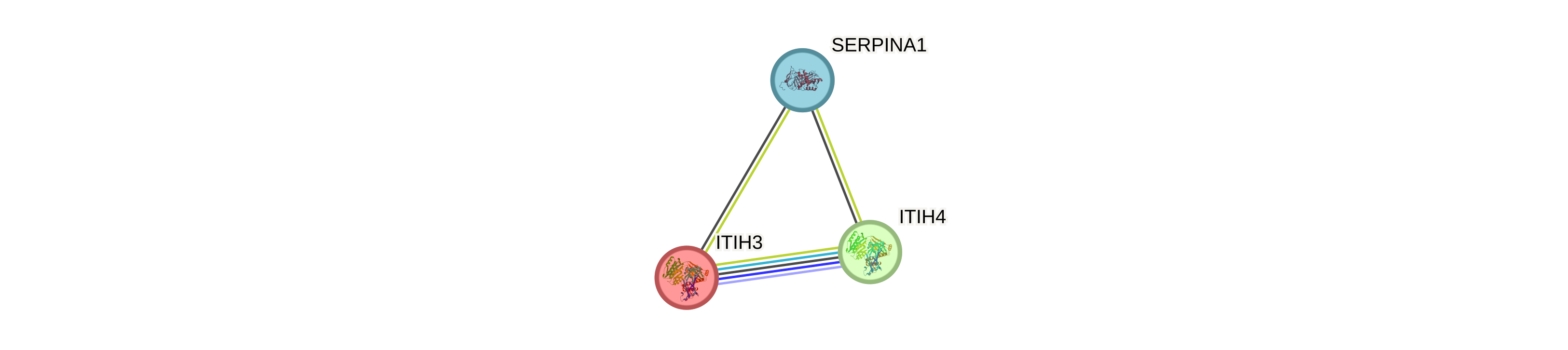

**Supplementary Figure 1.** StringDB^1^ network of three proteins demonstrating putative causal associations with neuropsychiatric disorders. SERPINA1 (alpha-1-antitrypsin) and ITIH4 (inter-alpha-trypsin inhibitor heavy chain 4) associated with autism-spectrum disorder. ITIH3 (inter-alpha-trypsin inhibitor heavy chain 3) associated with schizophrenia. Default StringDB settings were used: minimum required interaction score = 0.4, no limit to maximum number of interactors, edges represent protein-protein associations, light green = textmining, black = co-expression, light purple = protein homology, dark blue = gene co-occurrence, green = gene-neighbourhood. Download date: 05/03/2025.
